# Genomic epidemiology of *Klebsiella pneumoniae* neonatal and infant sepsis in Kenyan hospitals

**DOI:** 10.1101/2025.10.30.25339129

**Authors:** Anne V. Amulele, Benedict Orindi, Moreka Ndumba, Caroline Tigoi, Donwilliams Omuoyo, Eunice Kahindi, Wilson Gumbi, Leonard Ndwiga, Molly Timbwa, Christine Manyasi, Anne Kombo, Christina Obiero, Hermoine Webster, Nicole Stoesser, J. Anthony G. Scott, James A. Berkley

## Abstract

**Background:** *Klebsiella pneumoniae* (*Kpn*) is a leading cause of sepsis among hospitalised neonates and commonly causes outbreaks. Lack of surveillance data from low- and middle-income countries hampers development of vaccines and new measures to control infections. We describe the genomic epidemiology of *Kpn* neonatal infections admitted to Kenyan hospitals.

**Methods:** We analysed clinical and genome sequence data from neonates (≤28 days old) from a study of bacteraemia at admission at three Kenyan hospitals from October 2020 to April 2023 (NeoBAC), and, at one site, Kilifi, both neonates and infants (29–182 days old) from bacteraemia surveillance at admission and during admission from January 2001 to April 2023.

**Results:** We identified 75 neonatal cases of *Kpn* bacteraemia in NeoBAC, and 267 neonatal cases and 36 infant cases in the Kilifi surveillance. In NeoBAC, most neonatal infections were early onset and in-born whilst in the Kilifi surveillance most were late onset and out-born. Mortality was 32% (24/75) in NeoBAC, 41% (110/267) in Kilifi neonates and 56% (20/36) among Kilifi infants. Of 13 different STs identified in NeoBAC, ST6775 (29/75, 39%) and ST14 (26/75, 37%) were the most common. In Kilifi neonates 112 STs were identified, with ST17 (47/270, 17%) and ST14 (24/270, 9%) as the most common. Kilifi neonates had 61 KL- and 10 O-loci with predominant K-loci changing over the study period. Outbreak transmission clusters comprised 61/75 (81%) cases in NeoBAC and 140/270 (48%) cases in Kilifi neonates. Cumulative distribution suggested >30 K types would be needed in a vaccine to cover >85% of isolates.

**Conclusions:** *Kpn* bacteraemia occurred mainly in outbreaks with high mortality. The temporal dynamics of the target surface antigens may challenge future vaccine efficiency.

## INTRODUCTION

*Klebsiella pneumoniae* (*Kpn*), a Gram-negative bacterium, is a frequent cause of sepsis in neonates, the elderly and immunocompromised people (1). It is responsible for outbreaks in neonatal units, intensive care units, hospital wards and care homes (2–7). Globally, *Kpn* is a leading cause of infection-related (8) and antimicrobial resistance (AMR) related deaths (9).

Hospitalised preterm and low birth weight neonates are especially susceptible to *Kpn* sepsis because of extended hospital stay, medical devices, and immature immune and mucosal barrier functions (10). These factors, coupled with limited treatment options for multi-drug resistant (MDR) infections have recently resulted in prioritisation of *Kpn* for development of a maternal vaccination in pregnancy to protect infants (11). Such a vaccine could potentially avert ∼80,000 neonatal deaths annually worldwide (12), with the largest benefits in Africa and South Asia where the burden of neonatal sepsis and mortality is highest.

In Kenya, neonatal *Kpn* outbreaks have occasionally, but not systematically, been reported (3, 13, 14), and robust estimates of incidence, outcomes, and characteristics of invasive *Kpn* are lacking. We aimed to determine the frequency, genomic diversity, AMR profile, and timing of *Kpn* causing neonatal and early infant bacteraemia in three hospitals in Kenya.

## METHODS

### Study design, setting and participants

The study utilised isolates from NeoBAC, a prospective surveillance study of neonatal sepsis at three hospital sites in Kenya in 2020–23, and the long-term paediatric invasive bacterial disease surveillance at one site over two decades (15).

### The NeoBAC study

NeoBAC recruited neonates (≤28 days old) admitted at Kilifi County Teaching and Referral Hospital (KCTRH) serving the Kilifi Health and Demographic Surveillance System (16) and the wider Kilifi County on the Kenyan coast; Mbagathi County Hospital (MCH) in Nairobi; and Kiambu County Hospital (KCH) close to Nairobi. Between October 2020 and April 2023, neonates first starting intravenous antibiotics for suspected sepsis had blood cultures performed at that time. Subsequent blood cultures were not routinely available at two of the sites, so analysis of NeoBAC blood cultures from Kilifi were restricted to admission cultures, recognising that these overlapped with the Kilifi long term neonatal cases.

### The Kilifi long term surveillance

Paediatric invasive bacterial disease surveillance has been carried out at KCTRH for over two decades (15). All admissions to the paediatric wards were investigated with blood culture on admission except for elective surgery and minor injuries. Subsequent blood cultures were collected at clinicians’ discretion. We included isolates from neonates (≤28 days old) and infants to age 6 months (29–182 days old) admitted to KCTRH between January 2001 and April 2023 to characterise *Kpn* infections in and immediately beyond the neonatal period. Sequence data on isolates from January 2001 to December 2011 from Kilifi have previously been published (3).

### Ethical considerations

Ethical approval was provided by the KEMRI Scientific and Ethics Review Unit (SERU) and written informed consent was obtained from the parent/guardian of each participant (KEMRI SSC #1433, SERU #3957). Approval for the pooled analysis presented here was obtained from SERU (#4687).

### Laboratory methods

Blood cultures from MCH and KCH were transported to the KEMRI Centre for Microbiology Research (CMR) (17) in Nairobi and cultured in automated BACTEC incubators (BD, USA). Isolate species were identified using biochemical methods (API, bioMérieux, France). Samples from KCTRH were cultured in BACTEC incubators at the KEMRI-Wellcome Trust Research Programme (KWTRP)(18) were identified by API up to 2018 and thereafter by matrix-assisted laser desorption ionization-time of flight (MALDI-TOF, Bruker, Germany). Isolates from KCH and MCH were confirmed with MALDI-TOF at KWTRP. At both labs, *Kpn* isolates were stored at −80°C in tryptic soy broth with 15% glycerol.

### Antimicrobial resistance testing

Antimicrobial susceptibility testing (AST) was done by disc diffusion (Oxoid, UK). At KWTRP, breakpoints of the British Society of Antimicrobial Chemotherapy guidelines were followed initially, replaced in 2012 by those of the Clinical and Laboratory Standards Institute (CLSI). CLSI breakpoints were used throughout at CMR, Nairobi.

### Whole genome sequencing

Total DNA was extracted from single colony purity plates using QIAamp Fast DNA stool mini kit (Qiagen, UK) in Kilifi or Genomic tip 100/G (Qiagen, UK) at Modernising Medical Microbiology, University of Oxford. Library preparation was performed using the Illumina DNA Prep. Sequencing was performed on MiSeq (Illumina, USA) in Kilifi or the International Livestock Research Institute (Nairobi) producing 250 bp paired end reads. Sequencing was performed on NovaSeq 6000 (Illumina, USA) at Wellcome Trust Centre for Human Genomics, Oxford producing 150 bp paired end reads.

QC, genome assembly, genotyping and phylogenetic analysis

Quality control of raw reads was performed using FastQC v0.12.1 and filtering with FastP v0.23.4 set at 20 (19) removed low quality reads and adapter contamination. *De novo* assembly was done using SPAdes v3.15.5 (20). Assemblies that were 4.5–6.5 Mb, contigs ≤1000 and N50 >10,000 were included in the analysis.

Sequence types (ST), AMR and virulence genes were analysed using Kleborate v3.1.2 (21). Capsule (K) and lipopolysaccharide (LPS) (O) loci and antigen prediction was performed using Kaptive v3.0.0 (22).

Core genome single nucleotide polymorphisms (SNPs) were identified using snippy v4.6.0 at a minimum coverage of 5 (https://github.com/tseemann/snippy) by aligning raw reads against relevant reference genomes. For *K. pneumoniae* subsp. *pneumoniae* isolates NTUH-K2044 (AP006725.1) was used as the reference genome (23); for *K. quasipneumoniae*, ATCC 700603 (CP029597.1) (24); and for *K. variicola*, At-22 (CP001891.1) (25). Recombinant regions were filtered using gubbins v3.3.5 set at minimum SNP flag of 5 (26) and a maximum likelihood phylogeny inferred using RAxML (27). For each species grouping, SNP-only alignments were extracted using SNP-sites v2.5.1 (28) and pairwise genetic distances between the sequences were calculated using SNP-Dists v0.8.2 (https://github.com/tseemann/snp-dists) and IQ-TREE v2.3.4 (29) used for phylogenetic tree generations. Default parameters were used throughout unless otherwise specified.

### Statistical analysis

We summarised the characteristics of participants with *Kpn* infections across three groups: NeoBAC neonates, Kilifi neonates and Kilifi infants by demographic and baseline clinical characteristics, and by early or late onset of infection. All incidences or proportions were calculated on either NeoBAC or Kilifi data separately unless otherwise mentioned. Isolates from cultures taken up to 72 hours from birth were defined as early-onset infections (EOS), while those obtained after 72 hours were defined as late-onset infections (LOS). We estimated mortality during admission as a proportion with 95% Wilson’s confidence intervals. We studied associations between patient characteristics and mortality using logistic regression; characteristics significant in a univariable analysis at P<0.1 on likelihood ratio test (LRT) were included in a multivariable model. Duration of hospital stay was compared across groups using the Kruskal-Wallis test.

Diversity was summarised by richness (e.g., the number of unique STs) and abundance (e.g., the frequencies of a particular ST) and quantified using Simpson’s Diversity Index (D); D ranges from 0 (no diversity) to 1 (highest diversity). We summarised virulence and antibiotic resistance genes of strains isolated across groups. Outbreaks were investigated using clusters defined within a study site based on single-linkage clustering, using pairwise thresholds of genetic distance ≤5 SNPs and temporal distance ≤4 weeks, using <u>Transmission Estimator</u> <u>v0.1</u>(30). Transmission cluster analysis was performed for *K. pneumoniae* subsp. *pneumoniae* in the Kilifi surveillance. Analyses were performed using Stata software v17 (StataCorp, College Station, TX) and R v4.4.3 (R Core Team).

### Data availability

Sequence data generated in this study have been deposited in the National Centre for Biotechnology Information (NCBI) Short Reads Archive (SRA) under BioProject PRJNA1265413. Sequence data from isolates from 2001 to 2011 (3) are available from European Nucleotide Archive under project <u>PRJEB1563</u>. SRA genome accession numbers and associated metadata for the genomes are available at: https://doi.org/10.7910/DVN/0XINJB.

## RESULTS

### Participant characteristics

#### NeoBAC: Neonates

Blood cultures were performed in 8,558 neonates. *Kpn* was identified in 75 (0.88%) neonates; 43 (57%) were female and 62 (73%) were born in the study hospital (Table 1). Among the 75,

**Table 1.** Participant characteristics of neonates and infants with *K. pneumoniae* isolates that were sequenced^‡^

| Characteristic | NeoBAC neonates |  |  |  | Kilifi surveillance neonates |  |  |  | Kilifi surveillance infants |
| --- | --- | --- | --- | --- | --- | --- | --- | --- | --- |
|  | All | Early onset | Late onset | P-value | All | Early onset | Late onset | P-value | All |
| All | 75 | 60 (80) | 15 (20) | <0.001 | 267 | 57 (21) | 210 (79) | <0.001 | 36 |
| Hospital and County, no. (%)<sup>§</sup> |  |  |  | <0.001 |  |  |  |  |  |
| KCTRH in Kilifi | 9 | 4 (44) | 5 (56) |  | 267 | 57 (21) | 210 (79) |  | 36 |
| KCH in Kiambu | 46 | 44 (96) | 2 (4) |  | - | - | - |  | - |
| MCH in Nairobi | 20 | 12 (60) | 8 (40) |  | - | - | - |  | - |
| Year, no. (%) |  |  |  |  |  |  |  | <0.001 |  |
| 2001–05 | - | - | - |  | 14 | 5 (36) | 9 (64) |  | 12 |
| 2006–10 | - | - | - |  | 36 | 1 (3) | 35 (97) |  | 9 |
| 2011–15 | - | - | - |  | 95 | 15 (16) | 80 (84) |  | 7 |
| 2016–20 | - | - | - |  | 98 | 33 (34) | 65 (66) |  | 4 |
| 2021–23 | - | - | - |  | 24 | 3 (13) | 21 (88) |  | 4 |
| Admission age, median (IQR) days | 0 (0–0) | 0 (0–0) | 0 (0–15) | 0.004 | 0 (0–2) | 0 (0–1) | 0 (0–3) | 0.044 | 71 (40–93) |
| Sex female, no. (%) | 43 | 35 (81) | 8 (19) | 0.726 | 132 | 29 (22) | 103 (78) | 0.806 | 10 |
| Born at study hospital, no. (%) | 62 | 51 (82) | 11 (18) | 0.286 | 106 | 28 (26) | 78 (74) | 0.107 | 5 |
| Weight categories, no. (%)<sup>*</sup> |  |  |  | 0.055 |  |  |  | 0.077 |  |
| Very low (<1.5 kg) | 15 | 9 (60) | 6 (40) |  | 124 | 20 (16) | 104 (84) |  | 2 |
| Low (1.5 to <2.5 kg) | 28 | 22 (79) | 6 (21) |  | 94 | 23 (24) | 71 (76) |  | 4 |
| Normal ($\geq 2.5$ kg) | 31 | 28 (90) | 3 (10) | | 41 | 13 (32) | 28 (68) | | 27 |
| Missing | 1 | 1 (100) | 0 |  | 8 | 1 (13) | 7 (88) |  | 3 |
| Premature, no. (%) | 31 | 29 (94) | 2 (6) | 0.532 | 160 | 31 (19) | 129 (81) | 0.365 |  |
| Length of stay, median (IQR) days<sup>†</sup> | 10 (5–19) | 10 (5–16) | 15 (5–25) | 0.201 | 15 (7–27) | 6 (3–22) | 17 (8–30) | <0.001 | 14 (7–18) |
| Died in hospital, no. (%) | 24 | 17 (71) | 7 (29) | 0.173 | 110 | 27 (25) | 83 (75) | 0.275 | 20 |

‡ Early onset neonatal infection was defined as a *Kpn* identified in samples collected from neonates up to 72 hours from birth, while late onset infection was after 72 hours.
\*Weight was either birth or admission weight.
§ KCTRH is Kilifi County Teaching and Referral hospital; MCH is Mbagathi County Hospital; KCH is Kiambu County Hospital
† Estimated for survivors only.

60 (80%) were EOS, 28 (37%) had low birth weight (LBW, <2.5kg), 15 (20%) had very low birth weight (VLBW, <1.5kg) and 41 (55%) were reported preterm. Nine (12%) of NeoBAC *Kpn* cases were identified in Kilifi.

#### Kilifi long term surveillance: Neonates

Blood cultures were performed in 20288 neonatal admissions and 297 *Kpn* was identified in 285 (1.4%) neonates. Of 290 extracted and sequenced isolates, two were non-*Kpn*, seven were from later blood cultures with the same ST as the initial blood culture, and eleven failed the set quality control thresholds. In three neonates, a different ST was observed in later blood cultures from the initial blood culture and isolates from both timepoints were included. Thus, we included 270 non-duplicate *Kpn* isolates from 267 neonates in this analysis.

Of the 267 neonates with invasive *Kpn*, 132 (49%) were female, 106 (40%) were born in KCTRH, 57 (21%) were EOS, 94 (35%) were LBW, 124 (46%) were VLBW and 160 (60%) were reported preterm (Table 1).

#### Kilifi long term surveillance: Infants

Blood cultures were performed in 10685 admissions aged 29–182 days and 42 *Kpn* were identified in 41 (0.38%) infants. Of 41 extracted and sequenced isolates, five failed the set quality control thresholds and 36 non-duplicate *Kpn* were included in the analysis. Ten (28%) infants were female and the median age at admission was 71 days (IQR 40–93, Table 1).

### Mortality and duration of hospital stay

In NeoBAC neonates, 24/75 (32%, 95% CI 22–43%) died during admission compared with 110 (41%, 95% CI 35–47%) of 267 Kilifi neonates, and 20 (56%, 95% CI 40–70%) of 36 Kilifi infants (Table 1). Impatient mortality was positively associated with LBW and VLBW (S1 Table). The median length of hospital stay for survivors was 21 days overall, 10 days in NeoBAC neonates, 14 days in Kilifi infants and 15 days in Kilifi neonates (Table 1).

### K. pneumoniae diversity

#### Sequence types

Overall, we identified 136 unique STs, seven of which were novel: ST6586, ST6774, ST6775, ST6776, ST6777, ST6789 and ST6790. The STs were less diverse in NeoBAC (D=0.620) than in Kilifi neonates (D=0.952) and infants (D=0.967).

In NeoBAC samples 13 different STs were identified; ST6775 and ST14 accounted for 76% (57/75) of isolates (Fig 1, S2 Table). ST6775, was a novel ST observed only in neonates in KCH (n=29) between December 2021 and February 2022, consistent with a local outbreak. ST14, a known global “high-risk” lineage, was isolated from 13 neonates at KCH (July–August 2022) and from 13 neonates at MCH (September–October 2022).

**Fig 1.**
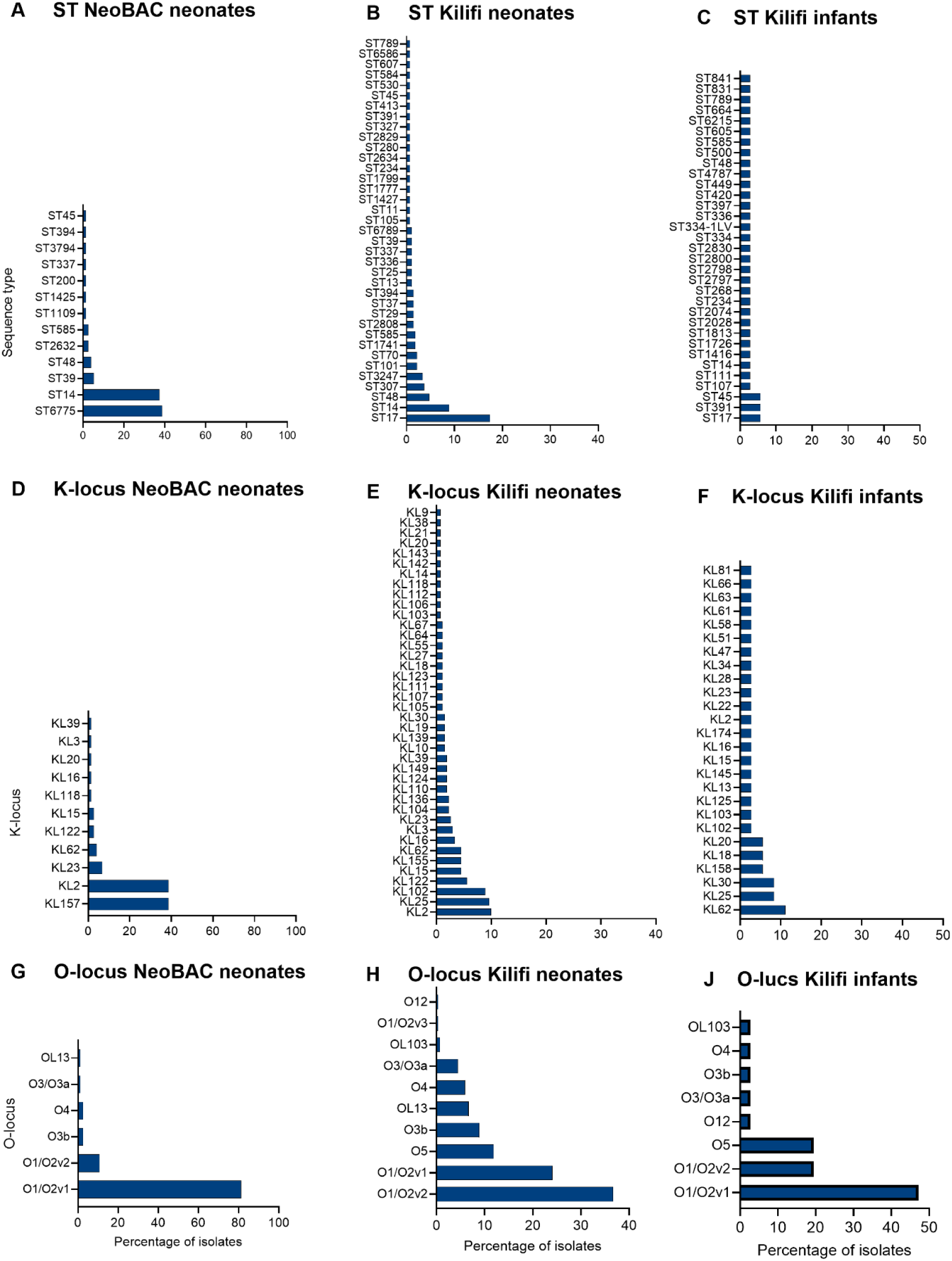
Relative abundance of main sequence types (ST) and surface antigen locus in *K. pneumoniae* isolates. The distribution of the ST (top panels), K–locus (middle panels) and O– locus (bottom panels) in hospitalised NeoBAC neonates (column 1, n=75 isolates) Kilifi neonates (column 2, n=270 isolates) and Kilifi infants (column 3, n=36).

Among Kilifi neonates, we observed 112 unique STs most commonly ST17 and ST14 which occurred in clusters over the years (Fig 1, Fig 2, S2 Table). Predominant STs changed over the study period. Nine cases due to ST3247 were in December 2017–January 2018 only. Among 36 isolates of *Kpn* detected in infants in Kilifi, there were 33 different STs (Fig 1, S2 Table).

**Fig 2.**
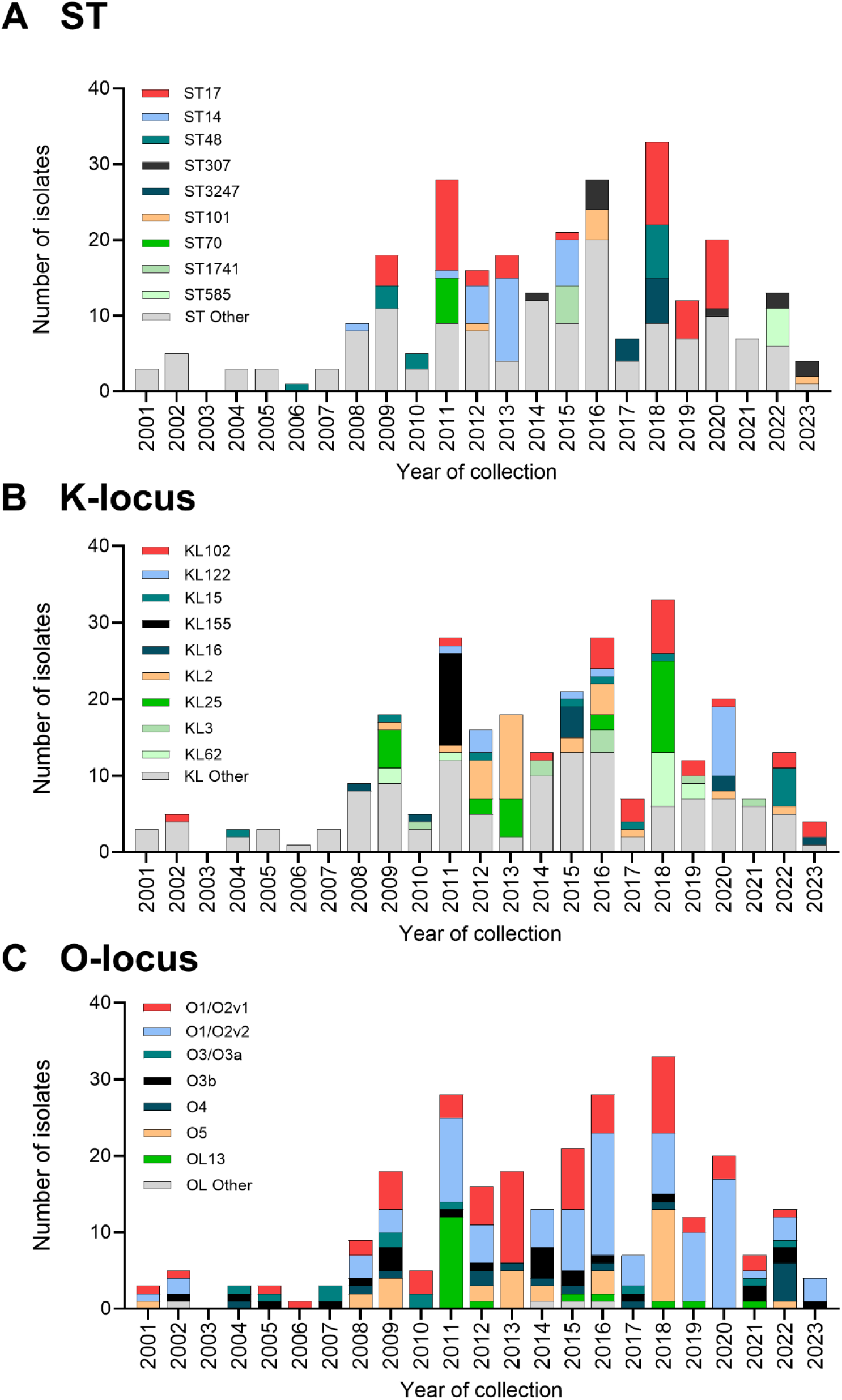
Distribution of main STs and surface antigen locus of *K. pneumoniae* isolates over the sampling years in the Kilifi neonatal population. The top 9 STs (of frequency ≥5) (A), 9 KL (of frequency ≥8) (B), and 7 OL (≥12 counts) (C) are shown individually, and the rest are grouped as other.

#### K and O antigen diversity

In NeoBAC, 11 unique KL types were identified. KL157 and KL2 were each detected in 29/75 (39%) cases, while the LPS (O) antigen locus O1/O2v1 was present in 61/75 (81%) and O1/O2v2 in 8/75 (11%) isolates (Fig 1, Tables S3, S4). KL157/O1/O2v1 was unique to the novel ST6775.

Among isolates from Kilifi neonates, there were 61 different KL types. The leading capsular antigen locus was KL2 found in 27 (10%) of the isolates and followed by KL25, KL102 and KL122. The major O-antigen loci were O1/O2v2, O1/O2v1 and O5 found in 99 (37%), 65 (24%) and 32 (12%), respectively (Fig 1, Tables S3, S4). While predominant KL-type changed over the study period, the same O-loci maintained predominance (Fig 2). The cumulative frequencies for KL and OL types varied across the three populations, with Kilifi neonates showing the least steep curve such that if 30 KL or 5 OL types are incorporated in a vaccine formulation, 85% of the isolates from Kilifi neonates will be covered (S1 Fig). Among Kilifi infants, 26 different KL types and 8 OL types were detected (Fig 1, Tables S3, S4). The Simpson’s index for KL was 0.620 in NeoBAC, 0.956 in Kilifi neonates and 0.949 in infants. The OL diversity was very low in NeoBAC (D=0.165) but very high in Kilifi neonates (D=0.776) and infants (D=0.697).

#### Virulence genes

In NeoBAC, yersiniabactin (*ybt*, complete or truncated) was the only siderophore detected in 40/75 (53%) isolates, of which 32 had *ybt* 14/ICE*Kp5* yersinibactin pathogenicity island marker. No genes encoding for other known virulence markers, including colibactin, aerobactin and salmochelin nor hypervirulence (*rmpA2*) were found. In Kilifi neonates, *ybt* was found in 90 isolates (33%). Of the Kilifi infant isolates 14 (39%) exhibited *ybt* with *ybt* 10/ICE*Kp4* present in 5 (S5 Table).

#### Antimicrobial resistance phenotypes

In 75 NeoBAC isolates, resistance to gentamicin, amoxicillin-clavulanate and cefotaxime was found in 41(55%),70 (93%) and 70 (93%) respectively. In 270 Kilifi neonatal isolates, 198 (73%) were resistant to gentamicin,189 (74%) to amoxicillin-clavulanate and 219 (81%) to cefotaxime. In 36 Kilifi infant isolates, 22 (61%),16/27 (59%) and 20 (56%) were resistant to gentamicin, amoxicillin-clavulanate and cefotaxime respectively. Resistance to imipenem was in a single NeoBAC isolate and one Kilifi neonate isolate (Fig 3).

**Fig 3.**
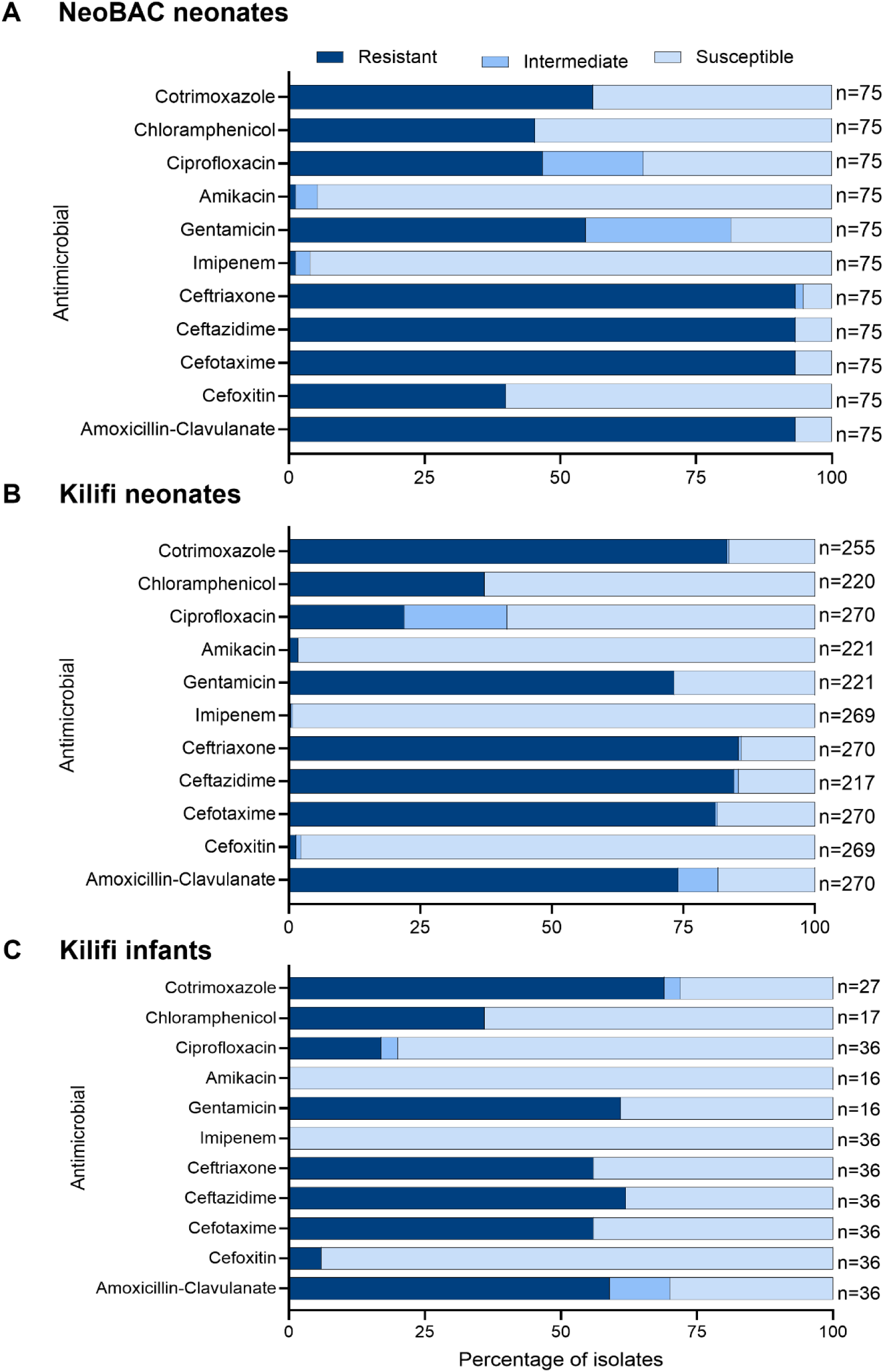
Antimicrobial susceptibility profile. *K. pneumoniae* isolates from NeoBAC neonates (A), Kilifi long-term surveillance neonates (B) and Kilifi long-term surveillance infants (C).

#### Antimicrobial resistance genotypes

In NeoBAC, the extended spectrum beta-lactamase gene (ESBL), *bla*CTX-M-15 was found in 67 of the 70 3GC resistant isolates. The *bla*NDM-5 carbapenemase was detected in the only imipenem-resistant isolate.

Among 270 isolates from Kilifi neonates in the long-term surveillance, 208 (77%) had *bla*CTX- M-15, 1 (0.5%) had *bla*CTX-M-14 and 60 (22%) had no ESBL gene; of these 10 (17%) were resistant to at least one 3GC on phenotypic testing. One isolate possessed *bla*NDM-5 and it was imipenem resistant. Among 36 isolates from Kilifi infants, 16 (44%) carried *bla*CTX-M-15 and of the remaining 20, four were resistant to at least one 3GC.

#### Outbreak and phylogenetic analysis

Outbreaks in neonatal units ranged in magnitude from a few cases lasting a few days to large outbreaks lasting several weeks. In NeoBAC, there were 6 clusters estimated using Transmission estimator, comprising 61 cases (81%, 95% CI 71–89%) and 5 STs, with a median cluster size of 8 (S2 Fig). The largest clusters were observed in KCH lasting 48 days in 2021– 2022 (29 isolates, ST6775) and 40 days in 2021 (13 isolates, ST14), and in MCH lasting 43 days in 2022 (13 isolates, ST14).

In Kilifi neonates, we estimated 29 clusters comprising 121 cases (49%, 95% CI 42–55%) and 19 STs, with a median cluster size of 3 (S3 Fig). The largest clusters lasted 37 days in 2017 (12 isolates, ST17) and 22 days in 2018 (12 isolates, ST17/ST336); the longest duration within a cluster was 39 days in 2017–2018 (9 isolates, ST3247). Among Kilifi infants there was a single cluster comprising two isolates in 2020 (ST45).

Consistent with our outbreak analyses, within the *K. pneumoniae* subsp. *pneumoniae* phylogenies we observed several closely related clusters representative of highly clonal lineages (SNP distribution range of 0–12) (Fig 4); this was similarly observed for *K. quasipneumoniae* (S4 Fig) and *K. variicola* (S5 Fig). Among ST17 isolates, three monophyletic clades were observed representing the main K-antigen types within ST17 (KL25, KL155 and KL122). Interestingly, four ST336 strains which also possessed KL25 antigen clustered together with the rest of the KL25 isolates of ST17. Clades of ST14 segregated by region; in Kilifi a major cluster represented KL2 antigen isolates and a minor cluster KL16. ST14 isolates from KCH and MCH formed a single clade though the isolates were from different years. ST14 isolates from MCH and KCH clustered within a single clade though the isolates were from different years.

**Fig 4.**
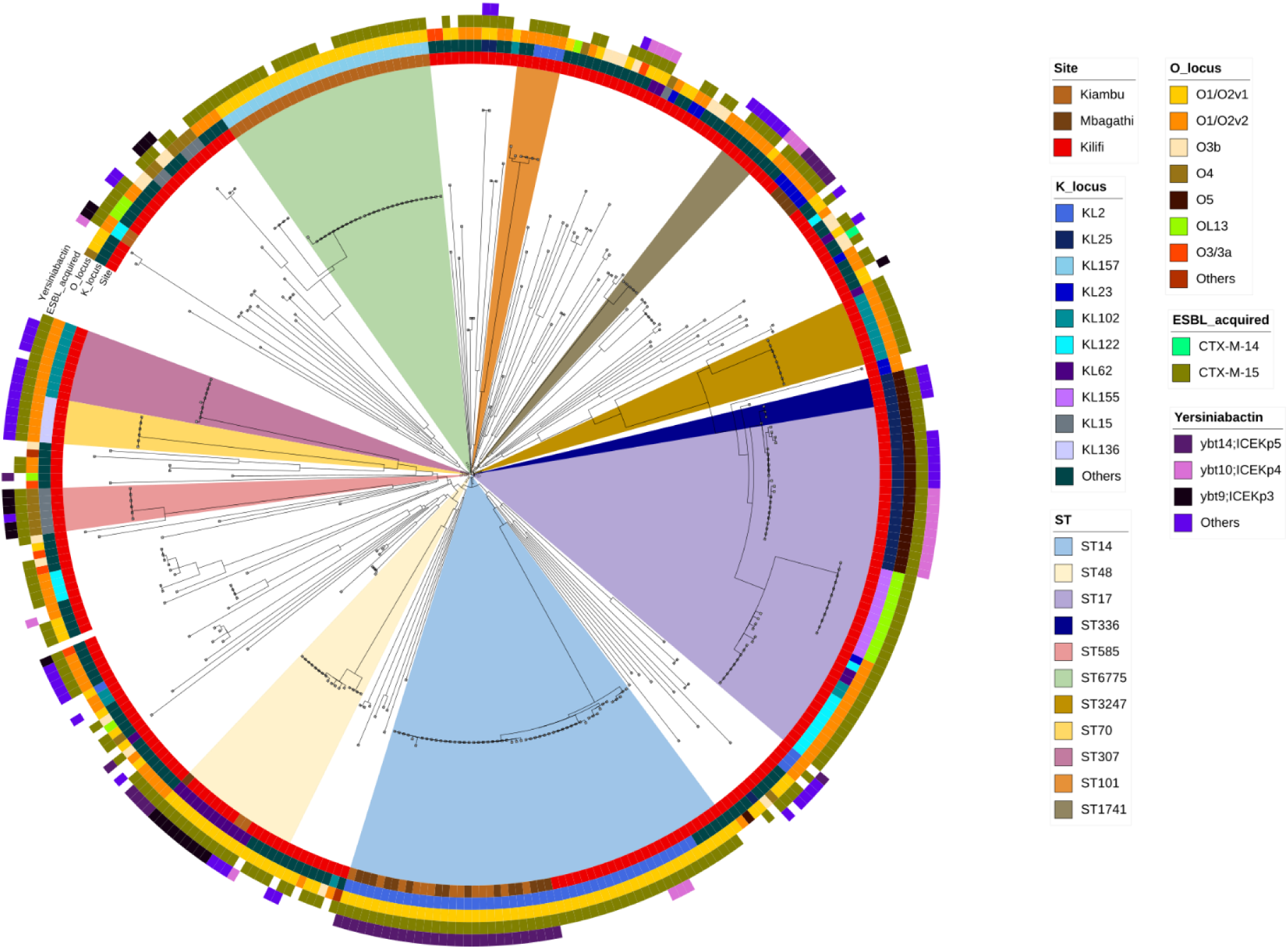
**Maximum likelihood phylogeny of 345 *K. pneumoniae* subsp. *pneumoniae* isolates**. Coloured rings (inner to outer) indicate site, K-antigen loci, O-antigen loci, ESBL genes (*bla*CTX-M only) and yersiniabactin. Colouring within the branches indicates the major ST isolated causing outbreaks within neonates. Distinct clades were observed within the dominant STs representing different K/O–loci in ST17 (KL155, Kl25, KL122) or different sites (ST14).

## DISCUSSION

We observed substantial diversity in the KL-types with fluctuating temporal strain dynamics and that outbreaks play an important role in KL-type dominance, raising questions about the durability of any K-antigen vaccine strategy. More than 30 KL-types would be needed to cover >85% of Kilifi neonatal cases compared to 5 OL-types for the same coverage. Our findings were similar to observations in Malawi, suggesting this might be a challenge across African settings (31). Some outbreak-associated KL-types (KL157, KL155) were unique to our setting and are therefore unlikely to be included in a vaccine, but may facilitate serotype replacement, a phenomenon observed for conjugate pneumococcal vaccines (32, 33).

Unsurprisingly given the sampling frame, the NeoBAC data showed predominance of early onset *Kpn* sepsis (60/75, 80%) among mostly inborn neonates. This contrasted with the longer-term picture revealed by the Kilifi surveillance data which showed predominance of late onset infections (210/267, 79%). These findings are consistent with the increasing prevalence of *Kpn* in sepsis occurring the first 72 hours of life (34). In BARNARDS, >50% of the *Kpn* infections across multiple low- and middle-income countries (LMICs) were late onset however, these included infants aged up to 60 days (35).

There was widespread resistance to first line antibiotics (amoxicillin–clavulanic acid/gentamicin) despite the fact that it was the most commonly used antibiotic in hospitalised neonates in Kenya (36). Resistance to 3GC was due to *bla*CTX-M-15 as was the case in other African and LMIC studies (31, 35). Resistance to carbapenems was rare in our study, as in Malawi (31) reflecting minimal use in government hospitals and suggesting a need for ongoing surveillance to monitor trends in resistance to improve carbapenem stewardship.

Outbreaks due to *Kpn* in neonatal units have been reported globally (2, 4, 6, 37, 38) and have been controlled by implementing measures such as improving hand hygiene practices, deep cleaning of hospital wards, limitation of ward traffic, reducing cot sharing and use of single use oxygen delivery devices, health education awareness to mothers, active surveillance cultures of patients and hospital environments (4, 6, 31, 39). Our findings suggest that the *Kpn* neonatal infection burden in Kenya is driven by hospital outbreaks, and improving hospital infection prevention control measures is a priority. The lack of microbiological investigations in many hospitals (40) suggests that many hospital outbreaks may be undetected and the burden of associated neonatal morbidity and mortality underestimated. This, coupled with evidence of interhospital transmission between MCH and KCH of ST14, which resulted in outbreaks in both hospitals in separate years provides a compelling reason for integrated surveillance across the hospital network.

In conclusion, a large proportion of *Kpn* sepsis cases in our study occurred within neonatal unit outbreaks. The temporal fluctuations of the diverse KL types observed in Kilifi have implications for future vaccine development. There was substantial resistance to recommended first line and second line treatment options though resistance to carbapenems was rare indicating a need for continued surveillance.

## ACKNOWLEDGEMENTS

We would like to acknowledge and thank the patients for their contribution to the studies, the clinical and laboratory staff in the collection of the samples, processing and storage of the isolates. We thank the International Livestock Research Institute (ILRI, Nairobi) for their support in sequencing and Adolf Mukama for computational support. We thank Erkison Odih and Kathyrn Holt for sharing the Transmission Estimator tool pre-publication.

## AUTHORS’ CONTRIBUTIONS

Conceptualization: AVA, JAB; investigation: MN, CT, HW, DO, WG, LN; data curation; AVA, MN, CT, DO, EK, WG, MT, AK, CM, CO; formal analysis: AVA, BO, MN; software: BO, MN, DO, WG; visualization: AVA, BO, MN; resources: CT, JAGS, NS, JAB; funding acquisition: CT, JAB; supervision: JAB; writing–original draft: AVA. All authors read and approved the final manuscript.

## SUPPORTING INFORMATION

**S1 Fig.**
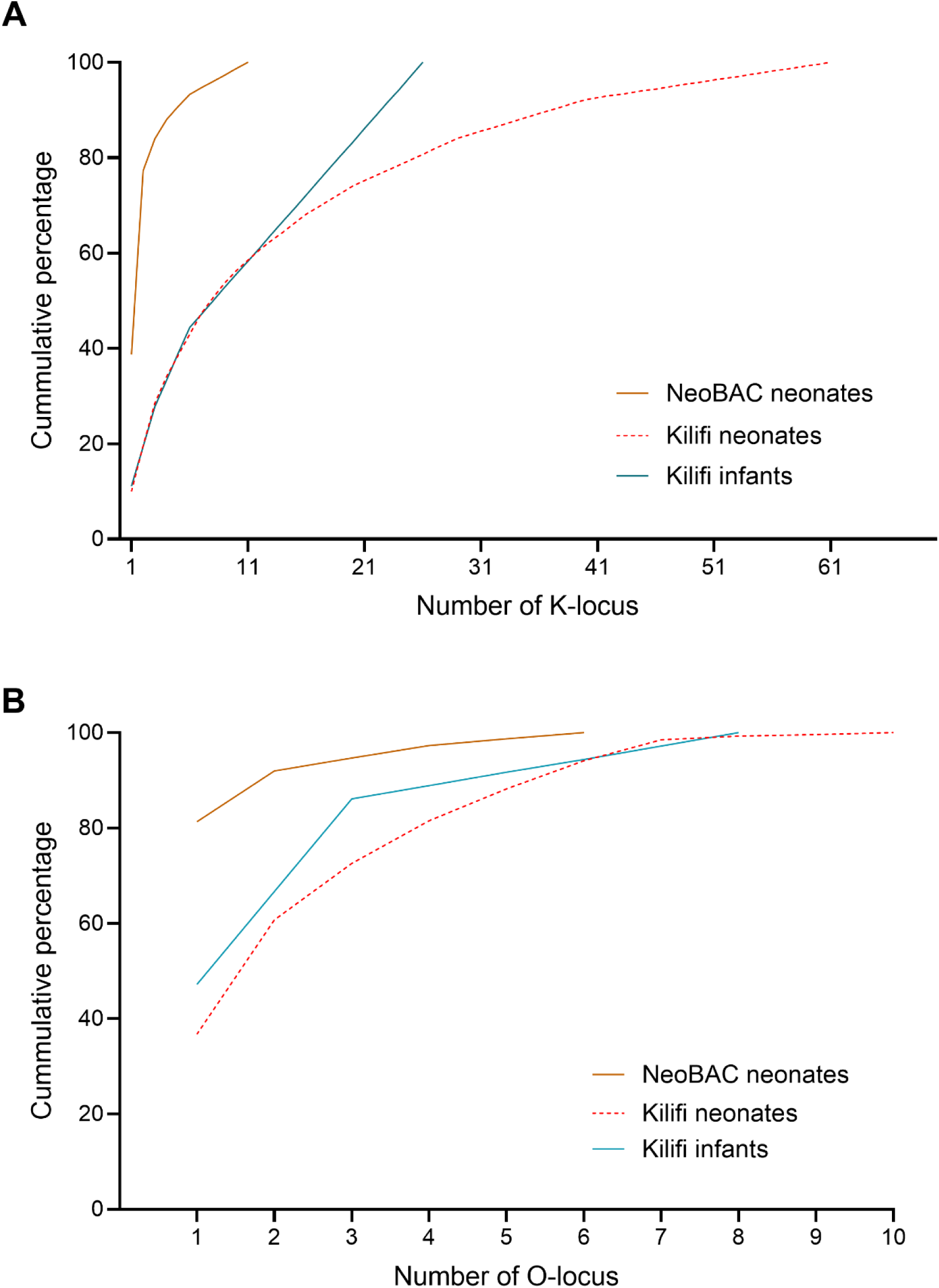
The cumulative coverage of the KL – types and the OL – types representing the percentage of isolates covered. The KL (A) and OL (B) types are sorted from the commonest.

**S2 Fig.**
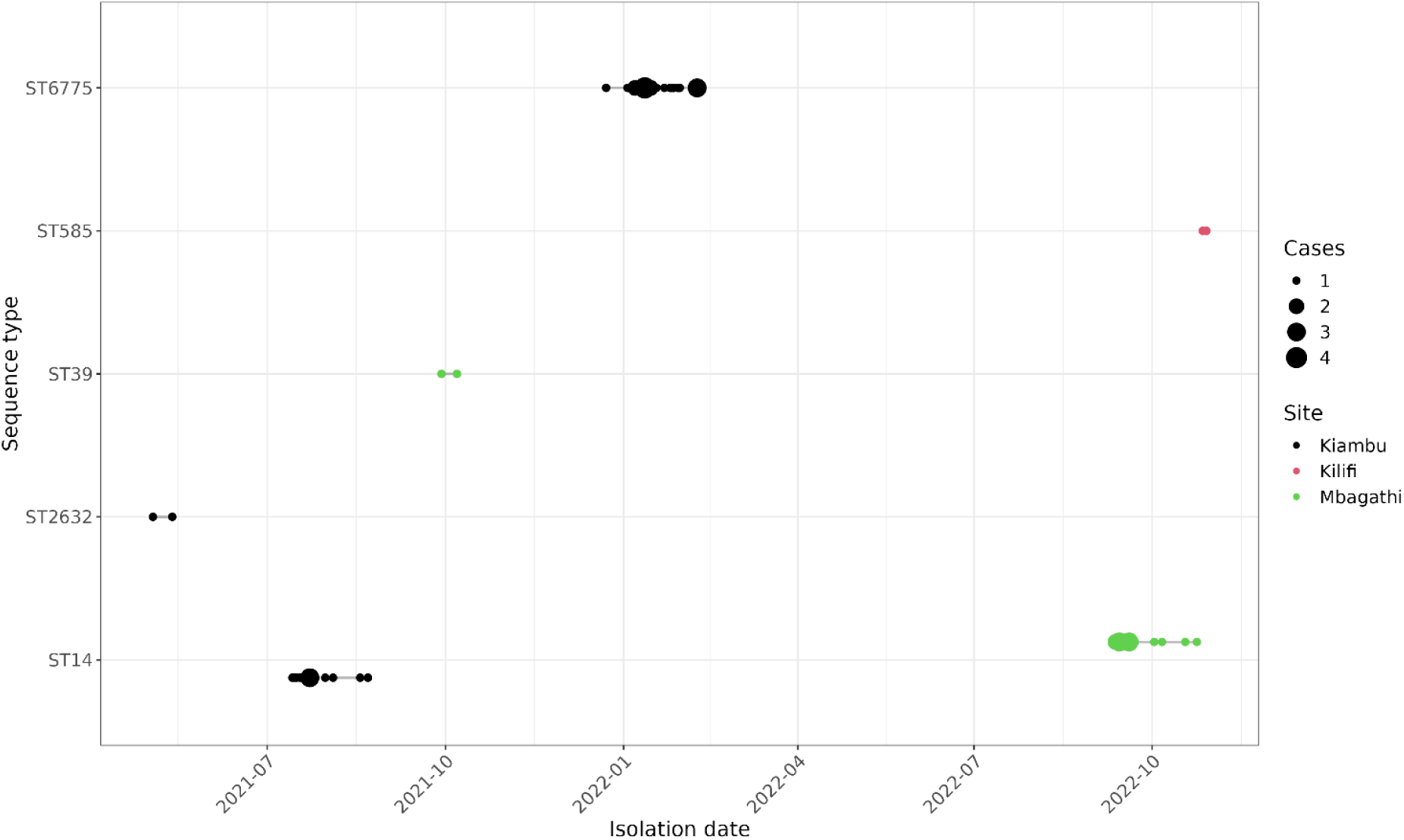
K. pneumoniae outbreaks within the NeoBAC population. Outbreaks were determined using Transmission Estimator v0.1 with a pairwise SNP distance threshold of ≤5 and temporal threshold of ≤4 weeks. Each point represents one or more cases isolated on specific dates. Clusters are represented as groups of cases (points) linked by horizontal lines. Sixty-one cases (81%) accounted for transmission clusters observed within sequence types (STs) and hospitals (KCH (black), MCH (green) and KCTRH (red)).

**S3 Fig.**
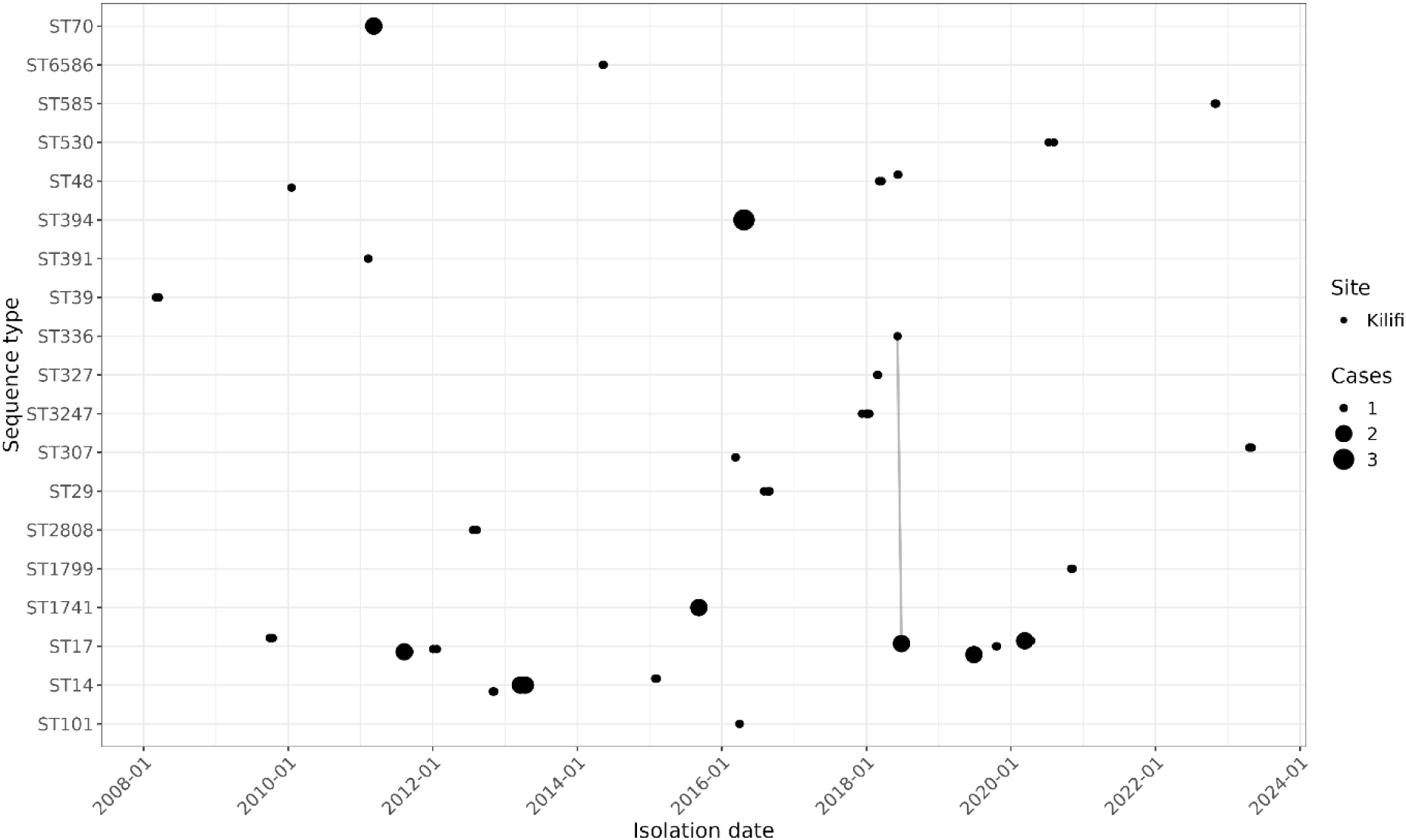
K. pneumoniae outbreaks within the Kilifi neonates population. Each point represents one or more cases isolated on specific dates. Clusters are represented as groups of cases (points). Transmission clusters were observed in 121 cases (49%) in both larger and smaller outbreaks occurring over several years. Clusters of ST17 and ST14 were observed more than once during the study period. Clustering between ST17 and ST336 in 2018 (vertical line) shows the temporal relatedness of the isolates observed to be related in the phylogeny.

**S4 Fig.**
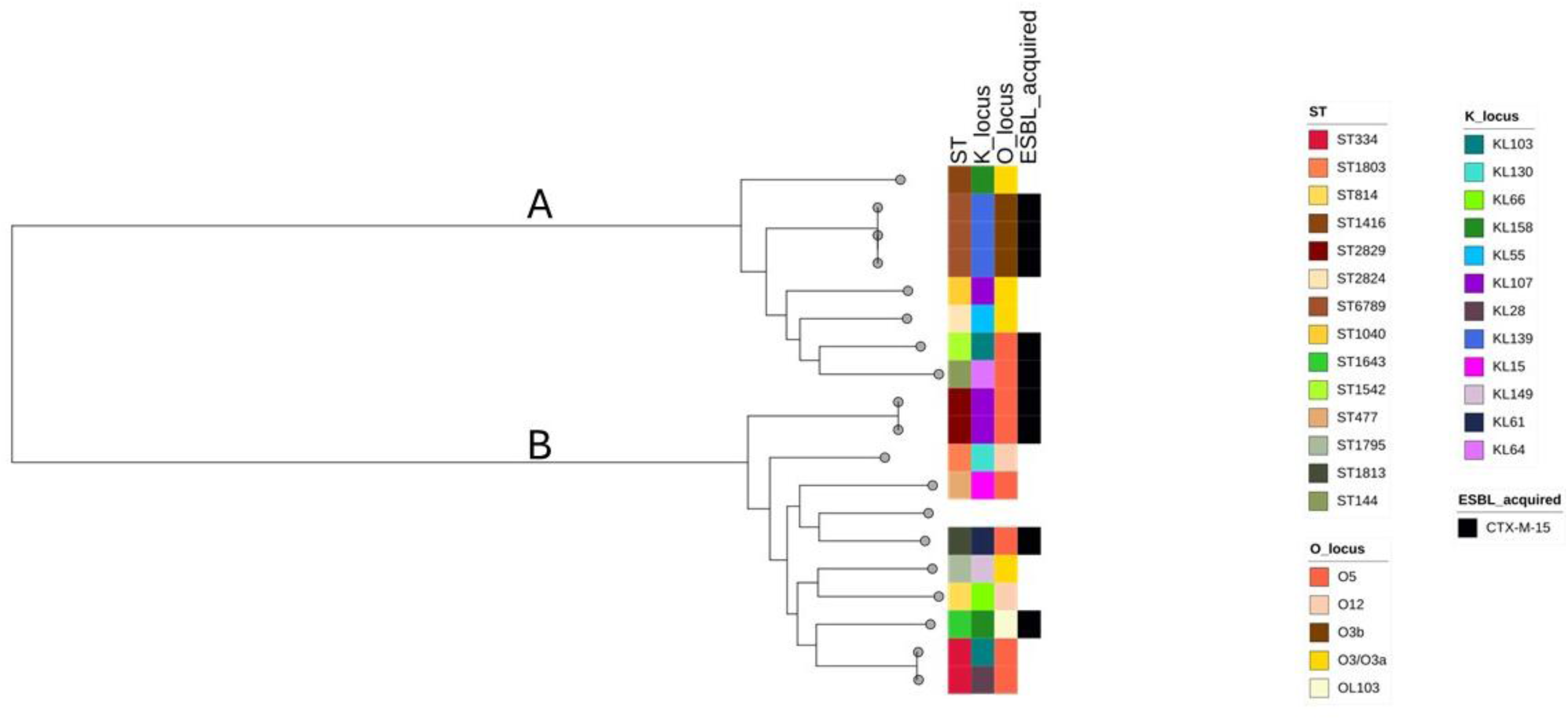
Phylogeny of 18 K. quasipneumoniae isolates. This subspecies was detected only in Kilifi comprising of eight K. quasipneumoniae subsp quasipneumoniae (A) and ten K. quasipneumoniae subsp similipneumoniae (B). Yersiniabactin genes were not detected in any of the strains.

**S5 Fig.**
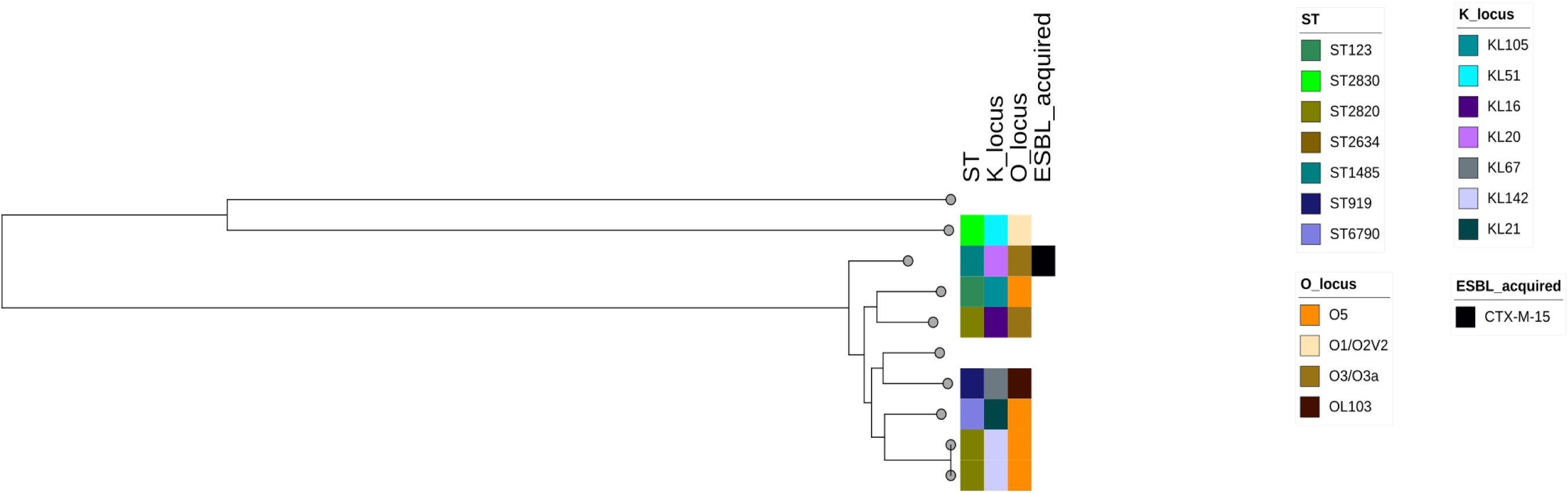
Phylogeny of 7 K. variicola and one K. quasivariicola isolates in Kilifi. Yersiniabactin genes were not detected in any of the strains

**S1 Table.** Association of patient characteristics with mortality

|  | Dead, n/N (% , 95%CI) | Unadjusted odds ratio (95% CI) | P-value | Adjusted odds ratio (95% CI) | P-value |
| --- | --- | --- | --- | --- | --- |
| Infection onset |  |  |  |  |  |
| Early onset | 42/113 (38%, 29–47%) | Reference |  | Reference |  |
| Late onset | 87/220 (40%, 34–47%) | 1.12 (0.7–1.78) | 0.648 | 0.63 (0.35–1.13) | 0.120 |
| Infant onset | 20/36 (56%, 40–70%) | 2.08 (0.97–4.46) | 0.059 | 2.71 (1.00–7.4) | 0.051 |
| Hospital and County§ |  |  |  |  |  |
| KCTRH in Kilifi | 130/303 (43%, 38–49%) | Reference |  | Reference |  |
| KCRH in Kiambu | 12/46 (26%, 16–40%) | 0.46 (0.23–0.92) | 0.028 | 0.65 (0.27–1.55) | 0.326 |
| MCRH in Nairobi | 7/20 (35%, 18–57%) | 0.70 (0.27–1.8) | 0.460 | 0.78 (0.28–2.18) | 0.639 |
| Year |  |  |  |  |  |
| 2001–05 | 11/26 (42%, 26–61%) | Reference |  |  |  |
| 2006–10 | 18/45 (40%, 27–55%) | 0.91 (0.34–2.42) | 0.849 |  |  |
| 2011–15 | 44/102 (43%, 34–53%) | 1.03 (0.43–2.47) | 0.939 |  |  |
| 2016–20 | 42/102 (43%, 34–53%) | 1.02 (0.43–2.45) | 0.960 |  |  |
| 2021–23 | 34/94 (36%, 27–46%) | 0.77 (0.32–1.87) | 0.568 |  |  |
| Sex |  |  |  |  |  |
| Male | 77/187 (42%, 35–49%) | Reference |  |  |  |
| Female | 72/182 (40%, 33–47%) | 0.92 (0.60–1.39) | 0.688 |  |  |
| Born at study hospital |  |  |  |  |  |
| No | 93/199 (47%, 41–54%) | Reference |  | Reference |  |
| Yes | 55/169 (33%, 26–40%) | 0.54 (0.35–0.83) | 0.005 | 0.65 (0.4–1.06) | 0.088 |
| Weight categories |  |  |  |  |  |
| Very low (<1.5 kg) | 64/139 (46%, 38–55%) | 2.11 (1.21–3.70) | 0.009 | 3.31 (1.61–6.8) | 0.001 |
| Low (1.5 to <2.5 kg) | 54/123 (44%, 36–53%) | 1.94 (1.09–3.44) | 0.023 | 3.07 (1.52–6.17) | 0.002 |
| Normal (≥2.5 kg) | 27/95 (29%, 21–39%) | Reference |  | Reference |  |
| Premature birth |  |  |  |  |  |
| No | 66/161 (41%, 34–49%) | Reference |  |  |  |
| Yes | 74/190 (40%, 33–47%) | 0.93 (0.61–1.43) | 0.751 |  |  |

§ KCTRH is Kilifi County Teaching and Referral hospital; MCH is Mbagathi County Hospital; KCH is Kiambu County Hospital

**S2 Table.** Sequence type distribution for all isolates S3 Table. K-locus distribution for all isolates

| NeoBAC neonates |  | Kilifi neonates |  |  |  |  |  | Kilifi infants |  |
| --- | --- | --- | --- | --- | --- | --- | --- | --- | --- |
| ST | No. (%) | ST | No. (%) | ST | No. (%) | ST | No. (%) | ST | No. (%) |
| ST6775 | 29 (39%) | ST17 | 47 (17%) | ST111 | 1 (0.4%) | ST2831-1LV | 1 (0.4%) | ST17 | 2 (6%) |
| ST14 | 28 (37%) | ST14 | 24 (9%) | ST123 | 1 (0.4%) | ST309 | 1 (0.4%) | ST391 | 2 (6%) |
| ST39 | 4 (5%) | ST48 | 13 (5%) | ST1296 | 1 (0.4%) | ST313 | 1 (0.4%) | ST45 | 2 (6%) |
| ST48 | 3 (4%) | ST307 | 10 (4%) | ST1307 | 1 (0.4%) | ST3140 | 1 (0.4%) | ST107 | 1 (3%) |
| ST2632 | 2 (3%) | ST3247 | 9 (3%) | ST1401 | 1 (0.4%) | ST323 | 1 (0.4%) | ST111 | 1 (3%) |
| ST585 | 2 (3%) | ST101 | 6 (2%) | ST1408 | 1 (0.4%) | ST33 | 1 (0.4%) | ST14 | 1 (3%) |
| ST1109 | 1 (1%) | ST70 | 6 (2%) | ST1425 | 1 (0.4%) | ST340 | 1 (0.4%) | ST1416 | 1 (3%) |
| ST1425 | 1 (1%) | ST1741 | 5 (2%) | ST144 | 1 (0.4%) | ST36 | 1 (0.4%) | ST1726 | 1 (3%) |
| ST200 | 1 (1%) | ST585 | 5 (2%) | ST147 | 1 (0.4%) | ST372 | 1 (0.4%) | ST1813 | 1 (3%) |
| ST337 | 1 (1%) | ST2808 | 4 (1%) | ST1485 | 1 (0.4%) | ST3794 | 1 (0.4%) | ST2028 | 1 (3%) |
| ST3794 | 1 (1%) | ST29 | 4 (1%) | ST152 | 1 (0.4%) | ST405 | 1 (0.4%) | ST2074 | 1 (3%) |
| ST394 | 1 (1%) | ST37 | 4 (1%) | ST1540 | 1 (0.4%) | ST469 | 1 (0.4%) | ST234 | 1 (3%) |
| ST45 | 1 (1%) | ST394 | 4 (1%) | ST1542 | 1 (0.4%) | ST477 | 1 (0.4%) | ST268 | 1 (3%) |
|  |  | ST13 | 3 (1%) | ST1552 | 1 (0.4%) | ST482 | 1 (0.4%) | ST2797 | 1 (3%) |
|  |  | ST25 | 3 (1%) | ST1557 | 1 (0.4%) | ST491 | 1 (0.4%) | ST2798 | 1 (3%) |
|  |  | ST336 | 3 (1%) | ST1643 | 1 (0.4%) | ST54 | 1 (0.4%) | ST2800 | 1 (3%) |
|  |  | ST337 | 3 (1%) | ST1786 | 1 (0.4%) | ST551 | 1 (0.4%) | ST2830 | 1 (3%) |
|  |  | ST39 | 3 (1%) | ST1795 | 1 (0.4%) | ST5561 | 1 (0.4%) | ST334 | 1 (3%) |
|  |  | ST6789 | 3 (1%) | ST18 | 1 (0.4%) | ST560 | 1 (0.4%) | ST334-1LV | 1 (3%) |
|  |  | ST105 | 2 (1%) | ST1800 | 1 (0.4%) | ST610 | 1 (0.4%) | ST336 | 1 (3%) |
|  |  | ST11 | 2 (1%) | ST1803 | 1 (0.4%) | ST628 | 1 (0.4%) | ST397 | 1 (3%) |
|  |  | ST1427 | 2 (1%) | ST1958 | 1 (0.4%) | ST661 | 1 (0.4%) | ST420 | 1 (3%) |
|  |  | ST1777 | 2 (1%) | ST1962 | 1 (0.4%) | ST6774 | 1 (0.4%) | ST449 | 1 (3%) |
|  |  | ST1799 | 2 (1%) | ST198 | 1 (0.4%) | ST6776 | 1 (0.4%) | ST4787 | 1 (3%) |
|  |  | ST234 | 2 (1%) | ST200 | 1 (0.4%) | ST6788 | 1 (0.4%) | ST48 | 1 (3%) |
|  |  | ST2634 | 2 (1%) | ST22 | 1 (0.4%) | ST6790 | 1 (0.4%) | ST500 | 1 (3%) |
|  |  | ST280 | 2 (1%) | ST247 | 1 (0.4%) | ST716 | 1 (0.4%) | ST585 | 1 (3%) |
|  |  | ST2829 | 2 (1%) | ST2616 | 1 (0.4%) | ST788 | 1 (0.4%) | ST605 | 1 (3%) |
|  |  | ST327 | 2 (1%) | ST2703 | 1 (0.4%) | ST834 | 1 (0.4%) | ST6215 | 1 (3%) |
|  |  | ST391 | 2 (1%) | ST2800 | 1 (0.4%) | ST919 | 1 (0.4%) | ST664 | 1 (3%) |
|  |  | ST413 | 2 (1%) | ST2805 | 1 (0.4%) | ST985 | 1 (0.4%) | ST789 | 1 (3%) |
|  |  | ST45 | 2 (1%) | ST2806 | 1 (0.4%) | ST987 | 1 (0.4%) | ST831 | 1 (3%) |
|  |  | ST530 | 2 (1%) | ST2807 | 1 (0.4%) |  |  | ST841 | 1 (3%) |
|  |  | ST584 | 2 (1%) | ST281 | 1 (0.4%) |  |  |  |  |
|  |  | ST607 | 2 (1%) | ST2812 | 1 (0.4%) |  |  |  |  |
|  |  | ST6586 | 2 (1%) | ST2814 | 1 (0.4%) |  |  |  |  |
|  |  | ST789 | 2 (1%) | ST2815 | 1 (0.4%) |  |  |  |  |
|  |  | ST1026 | 1 (0.4%) | ST2817 | 1 (0.4%) |  |  |  |  |
|  |  | ST1040 | 1 (0.4%) | ST2820 | 1 (0.4%) |  |  |  |  |
|  |  | ST1109 | 1 (0.4%) | ST2824 | 1 (0.4%) |  |  |  |  |

**S3 Table.** K-locus distribution for all isolates

| NeoBAC neonates |  | Kilifi neonates |  |  |  | Kilifi infants |  |
| --- | --- | --- | --- | --- | --- | --- | --- |
| K-locus | No. (%) | K-locus | No. (%) | K-locus | No. (%) | K-locus | No. (%) |
| KL157 | 29(39%) | KL2 | 27 (10%) | KL21 | 2 (0.7%) | KL62 | 4 (11%) |
| KL2 | 29(39%) | KL25 | 26 (10%) | KL38 | 2 (0.7%) | KL25 | 3 (8%) |
| KL23 | 5(7%) | KL102 | 24 (9%) | KL9 | 2 (0.7%) | KL30 | 3 (8%) |
| KL62 | 3(4%) | KL122 | 15 (6%) | KL109 | 1 (0.4%) | KL158 | 2 (6%) |
| KL122 | 2(3%) | KL15 | 12 (4%) | KL125 | 1 (0.4%) | KL18 | 2 (6%) |
| KL15 | 2(3%) | KL155 | 12 (4%) | KL127 | 1 (0.4%) | KL20 | 2 (6%) |
| KL118 | 1(1%) | KL62 | 12 (4%) | KL130 | 1 (0.4%) | KL102 | 1 (3%) |
| KL16 | 1(1%) | KL16 | 9 (3%) | KL141 | 1 (0.4%) | KL103 | 1 (3%) |
| KL20 | 1(1%) | KL3 | 8 (3%) | KL151 | 1 (0.4%) | KL125 | 1 (3%) |
| KL3 | 1(1%) | KL23 | 7 (3%) | KL158 | 1 (0.4%) | KL13 | 1 (3%) |
| KL39 | 1(1%) | KL104 | 6 (2%) | KL163 | 1 (0.4%) | KL145 | 1 (3%) |
|  |  | KL136 | 6 (2%) | KL180 | 1 (0.4%) | KL15 | 1 (3%) |
|  |  | KL110 | 5 (2%) | KL24 | 1 (0.4%) | KL16 | 1 (3%) |
|  |  | KL124 | 5 (2%) | KL31 | 1 (0.4%) | KL174 | 1 (3%) |
|  |  | KL149 | 5 (2%) | KL42 | 1 (0.4%) | KL2 | 1 (3%) |
|  |  | KL39 | 5 (2%) | KL43 | 1 (0.4%) | KL22 | 1 (3%) |
|  |  | KL10 | 4 (1%) | KL46 | 1 (0.4%) | KL23 | 1 (3%) |
|  |  | KL139 | 4 (1%) | KL47 | 1 (0.4%) | KL28 | 1 (3%) |
|  |  | KL19 | 4 (1%) | KL49 | 1 (0.4%) | KL34 | 1 (3%) |
|  |  | KL30 | 4 (1%) | KL51 | 1 (0.4%) | KL47 | 1 (3%) |
|  |  | KL105 | 3 (1%) | KL54 | 1 (0.4%) | KL51 | 1 (3%) |
|  |  | KL107 | 3 (1%) | KL57 | 1 (0.4%) | KL58 | 1 (3%) |
|  |  | KL111 | 3 (1%) | KL61 | 1 (0.4%) | KL61 | 1 (3%) |
|  |  | KL123 | 3 (1%) | KL8 | 1 (0.4%) | KL63 | 1 (3%) |
|  |  | KL18 | 3 (1%) |  |  | KL66 | 1 (3%) |
|  |  | KL27 | 3 (1%) |  |  | KL81 | 1 (3%) |
|  |  | KL55 | 3 (1%) |  |  |  |  |
|  |  | KL64 | 3 (1%) |  |  |  |  |
|  |  | KL67 | 3 (1%) |  |  |  |  |
|  |  | KL103 | 2 (1%) |  |  |  |  |
|  |  | KL106 | 2 (1%) |  |  |  |  |
|  |  | KL112 | 2 (1%) |  |  |  |  |
|  |  | KL118 | 2 (1%) |  |  |  |  |
|  |  | KL14 | 2 (1%) |  |  |  |  |
|  |  | KL142 | 2 (1%) |  |  |  |  |
|  |  | KL143 | 2 (1%) |  |  |  |  |
|  |  | KL20 | 2 (1%) |  |  |  |  |

**S4 Table.** O-locus distribution for all isolates.

| NeoBAC neonates |  | Kilifi neonates |  | Kilifi infants |  |
| --- | --- | --- | --- | --- | --- |
| O-locus | No. (%) | O-locus | No. (%) | O-locus | No. (%) |
| O1/O2v1 | 61 (81%) | O1/O2v2 | 99 (37%) | O1/O2v1 | 17 (47%) |
| O1/O2v2 | 8 (11%) | O1/O2v1 | 65 (24%) | O1/O2v2 | 7 (19%) |
| O3b | 2 (3%) | O5 | 32 (12%) | O5 | 7 (19%) |
| O4 | 2 (3%) | O3b | 24 (9%) | O12 | 1 (3%) |
| O3/O3a | 1 (1%) | OL13 | 18 (7%) | O3/O3a | 1 (3%) |
| OL13 | 1 (1%) | O4 | 16 (6%) | O3b | 1 (3%) |
|  |  | O3/O3a | 12 (4%) | O4 | 1 (3%) |
|  |  | OL103 | 2 (1%) | OL103 | 1 (3%) |
|  |  | O1/O2v3 | 1 (0.4%) |  |  |
|  |  | O12 | 1 (0.4%) |  |  |

**S5 Table.** Frequencies of yersiniabactin for all isolates.

| NeoBAC neonates |  | Kilifi neonates |  | Kilifi infants |  |
| --- | --- | --- | --- | --- | --- |
| Yersiniabactin | No. (%) | Yersiniabactin | No. (%) | Yersiniabactin | No. (%) |
| *ybt* 14/ICEKp5 | 32 (43%) | *ybt* 10/ICEKp4 | 19 (7%) | *ybt* 10/ICEKp4 | 5 (14%) |
| *ybt* 9/ICEKp3 | 5 (7%) | *ybt* 9/ICEKp3 | 17 (6%) | *ybt* 14/ICEKp5 | 2 (6%) |
| *ybt* 14/ICEKp12 | 2 (3%) | *ybt* 14/ICEKp12 | 14 (5%) | *ybt* 15/ICEKp11 | 2 (6%) |
| *ybt* 10/ICEKp4 | 1 (1%) | *ybt* 16/ICEKp12 | 11 (4%) | *ybt* 9/ICEKp3 | 2 (6%) |
| *ybt* not detected | 35 (47%) | *ybt* 8/ICEKp9 | 11 (4%) | *ybt* 0/ICEKp10 | 1 (3%) |
|  |  | *ybt* 14/ICEKp5 | 6 (2%) | *ybt* 16/ICEKp12 | 1 (3%) |
|  |  | *ybt* 13/ICEKp2 | 5 (2%) | *ybt* 7/ICEKp7 | 1 (3%) |
|  |  | *ybt* 15/ICEKp11 | 3 (1%) | *ybt* not detected | 22 (61%) |
|  |  | *ybt* 6/ICEKp5 | 2 (1%) |  |  |
|  |  | *ybt* 0/ICEKp2 | 1 (0%) |  |  |
|  |  | *ybt* 22/ICEKp12 | 1 (0%) |  |  |
|  |  | *ybt* not detected | 180 (67%) |  |  |

